# A Multiomics Approach to Heterogeneity in Alzheimer’s Disease: Focused Review and Roadmap

**DOI:** 10.1101/19008615

**Authors:** AmanPreet Badhwar, G. Peggy McFall, Shraddha Sapkota, Sandra E. Black, Howard Chertkow, Simon Duchesne, Mario Masellis, Liang Li, Roger A. Dixon, Pierre Bellec

**Author notes:** **Corresponding Author** Dr. AmanPreet Badhwar, Centre de Recherche, Institut Universitaire de Gériatrie de Montréal, Université de Montréal, Montréal, QC, Canada H3W 1W5. equal contribution (Roger A. Dixon, Pierre Bellec).

## Abstract

Etiological and clinical heterogeneity is increasingly recognized as a common characteristic of Alzheimer’s disease and related dementias. This heterogeneity complicates diagnosis, treatment, and the design and testing of new drugs. An important line of research is discovery of multimodal biomarkers that will facilitate the targeting of subpopulations with homogeneous pathophysiological signatures. High-throughput ‘omics’ are unbiased data driven techniques that probe the complex etiology of Alzheimer’s disease from multiple levels (e.g. network, cellular, and molecular) and thereby account for pathophysiological heterogeneity in clinical populations. This review focuses on data reduction analyses that identify complementary disease-relevant perturbations for three omics techniques: neuroimaging-based subtypes, metabolomics-derived metabolite panels, and genomics-related polygenic risk scores. Neuroimaging can track accrued neurodegeneration and other sources of network impairments, metabolomics provides a global small-molecule snapshot that is sensitive to ongoing pathological processes, and genomics characterizes relatively invariant genetic risk factors representing key pathways associated with Alzheimer’s disease. Following this focused review, we present a roadmap for assembling these multiomics measurements into a diagnostic tool highly predictive of individual clinical trajectories, to further the goal of personalized medicine in Alzheimer’s disease.

## Introduction

Alzheimer’s disease is a complex, multifactorial pathology that manifests itself along a continuum of conditions, ranging from asymptomatic, to mild cognitive impairment (MCI), to dementia (specifically Alzheimer’s disease dementia). Trials of disease-modifying therapies remain unsuccessful, and these persistent failures have been attributed to (1) intervention late in the disease process (i.e. symptomatic stage), by which time extensive irreversible damage has accrued, and (2) lack of precision intervention targets in a multifactorial condition. Accordingly, an important line of current research is directed at the discovery of multimodal biomarkers that will help facilitate the detection of Alzheimer’s disease in asymptomatic populations, and the adaptation of intervention regimens to different target subpopulations in prevention trials (Anstey *et al*., 2015; Olanrewaju *et al*., 2015). This work reviews recent data-driven approaches to biomarker discovery, in three omics fields that capture complementary aspects of neurodegeneration and Alzheimer’s disease risk factors. We further propose a roadmap for integrating these multiomics biomarkers to advance our understanding of heterogeneity in Alzheimer’s disease, and promote efficacy in intervention trials.

Established Alzheimer’s disease biomarkers currently capture three facets of the disease pathophysiology: amyloidosis, tauopathy, and specific aspects of neurodegeneration (Jack *et al*., 2018). Although these biomarkers have been usefully applied to the crucial goal of early Alzheimer’s disease detection (Sperling *et al*., 2011), they fall short in explaining the heterogeneity of individual clinical trajectories, and their ability to predict differential cognitive decline is modest (Dumurgier *et al*., 2017). Predicting progression to dementia is challenging, as patients diagnosed with probable Alzheimer’s disease dementia show considerable heterogeneity in the cognitive domains impaired (Scheltens *et al*., 2016), and the presence or severity of established Alzheimer’s disease biomarkers. For example, amyloidosis-and-tauopathy-defined ‘pure Alzheimer’s disease neuropathology’ is observed in only 30-50 percent of patients with probable Alzheimer’s disease dementia (Beach *et al*., 2012; Robinson *et al*., 2018). The remaining cases show co-occurrence of multiple brain pathologies that overlap with other NDDs of aging, such as cerebral small vessel disease, and Lewy body dementia. Minimum to above-threshold levels of Alzheimer’s disease pathology are also observed in a considerable proportion (39%) of dementia patients not clinically diagnosed as probable Alzheimer’s disease (Beach *et al*., 2012). Alzheimer’s disease pathology has also been demonstrated in postmortem studies of CN older adults (Bennett *et al*., 2006), and it remains unclear if such individuals would have developed Alzheimer’s disease symptoms with time, should they have lived longer (Jagust, 2013). Overall, “top-down” clinical labels, based primarily on cognitive symptoms, imperfectly align with biomarkers of neurodegeneration. Additional biomarkers are thus urgently needed to characterize the clinico-pathological heterogeneity of Alzheimer’s disease, and to disambiguate it from other age-related NDDs and normal aging (Jack *et al*., 2018).

A radically different paradigm to NDDs is to move away from “top-down” clinical labels, and concentrate on pathological signatures built “bottom up”, using unsupervised machine learning algorithms and high-throughput ‘omics’ metrics that screen global facets of an organism. These data-driven approaches provide new opportunities to probe the complex etiology of Alzheimer’s disease from multiple levels (example network, cellular, and molecular), and to identify biomarker signatures with high diagnostic/prognostic value. This review focuses on the following omics approaches: brain connectomics, metabolomics, and genomics. These omics data capture complementary information on Alzheimer’s disease emergence and progression: brain connectomics (and morphometry) can track accrued neurodegeneration and other sources of network impairments, metabolomics provides a global small-molecule snapshot that is sensitive to ongoing pathological processes, and genomics characterizes relatively invariant genetic risk factors representing key pathways associated with Alzheimer’s disease (Jack *et al*., 2018). The high-dimensional nature of omics “big data” can prove challenging to process, manipulate, and visualize, even when a single modality is involved. Multiple redundancies are often present in these measures, and not all data points provide independent information as they tend to covary due to shared biological processes. We focus this review on three omics data reduction techniques that capture disease-relevant population heterogeneity with a limited number of indicators: neuroimaging-based subtypes, metabolite panels, and polygenic risk score or PRS. Neuroimaging subtypes are based on data-driven algorithms that identify patient subgroups with homogeneous brain imaging features. Metabolite panels are developed via data-driven algorithms applied to thousands of small molecules representing global biochemical events and distinguishing clinical phenotypes. PRS and other empirically derived representations of interactive or multi-gene risk may represent key domains of mechanisms and pathways to Alzheimer’s disease. Following this focused review, we discuss the rationale and challenges for assembling multiomics diagnostic tools highly predictive of individual clinical trajectories, and in particular, the importance of pathophysiological heterogeneity in research clinical cohorts.

## Materials and Methods

We conducted parallel focused reviews of PubMed articles published between January 2011 to June 2018 in three omics domains: brain connectomics (and morphometry), metabolomics, and genomics. We included studies investigating Alzheimer’s disease in human and published in English. Additional articles (that met criteria) were identified by scanning the reference lists of selected PubMed articles. Described in the following three sections are search characteristics specific to each omics domain. We have provided in Supplementary Material (Supplementary Tables S1, S2, and S5) characteristics of the 40 domain-specific omics studies included.

### Brain morphology and connectomics

Search term combinations used for brain morphology and connectomics in neuroimaging are provided in were: (1) alzheimer’s disease OR alzheimer pathology AND subtype, (2) mild cognitive impairment OR amnestic mild cognitive impairment OR MCI OR amnestic MCI AND hierarchical clustering; resting-state AND functional MRI AND alzheimer AND clustering, (3) alzheimer’s disease OR alzheimer pathology AND structural subtype, (4) alzheimer’s disease OR alzheimer pathology AND structural MRI AND clustering, (5) resting-state AND functional MRI AND alzheimer AND hierarchical clustering, (6) diffusion MRI AND alzheimer AND clustering, (7) diffusion MRI AND alzheimer AND hierarchical clustering; diffusion MRI AND hierarchical clustering. To these we applied the “common” exclusion criteria. Thereafter, only studies reporting AD spectrum subgroups identified using data-driven methods were included. It should be noted that neuroimaging-based subtyping is an emergent field and there is still a lack of consistent terminology. Therefore, to avoid missing relevant studies due to stringent use of terminology, search combinations 1 and 2, used relaxed and inclusive keywords, which captured all of the morphometry based literature. In total, 12 papers met our criteria, and were reviewed.

### Metabolomics

Search term combinations for metabolomics were: (1) alzheimer’s disease AND metabolomics panels; (2) alzheimer’s disease AND metabolomics profiles; (3) alzheimer’s disease AND metabolomics networks; (4) alzheimer’s disease AND metabolomics pathways; (5) alzheimer’s disease AND metabolomics biomarkers. To these we applied the common exclusion criteria. We further excluded: reviews and technical reports; articles not relevant to AD metabolomic panels, pathways, or networks; and studies using a targeted metabolomics approach. Five papers were identified from reference list scans. In total, 11 studies were reviewed. Details on key metabolites highted in the 11 studies were compiled from three different databases: Kyoto Encyclopedia of Genes and Genomes (KEGG, https://www.genome.jp/kegg/pathway.html) Pathway Database, Human Metabolome Database (HMDB, http://www.hmdb.ca/) and PubMed. These details were used to generate Fig. 2b and are provided in Supplementary Table S3.

**Figure 1:**
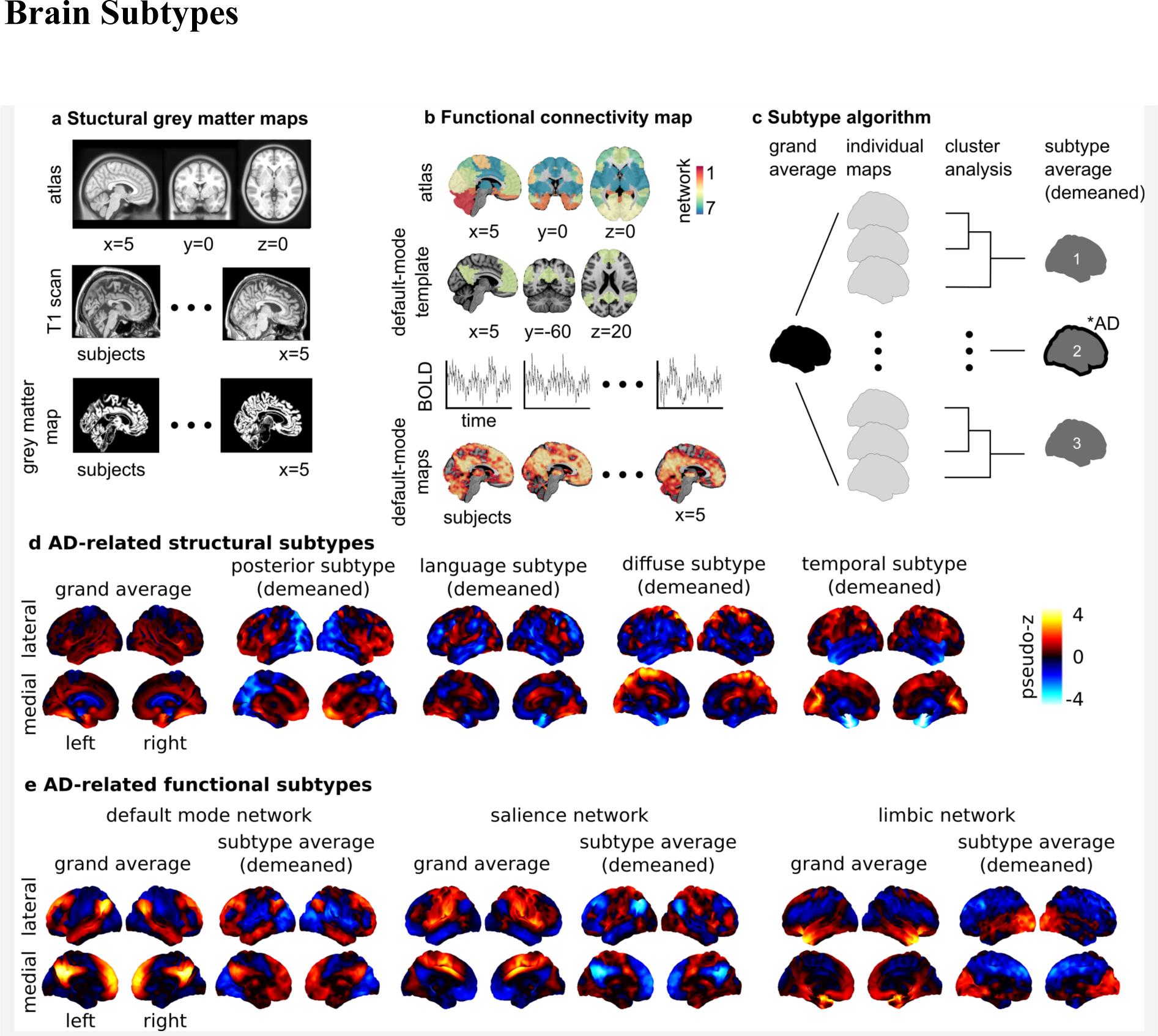
Brain morphology and connectomics Alzheimer’s disease-related subtypes. Neuroimaging provides insight into the effect of neurodegeneration on brain health. There exist different tools that can capture distinct, yet complementary, aspects of brain structure and function. The most established neuroimaging marker of neurodegeneration is grey matter atrophy, measured by structural MRI. Structural MRI is a non-invasive technique widely used in both research and clinical practice. To generate structural maps, individual structural MRI scans are first spatially aligned to a reference template or atlas (1a). Then for each individual and each voxel (smallest volume element in MRI data), a metric characterizing the local structure of the grey matter is generated (1a), such as grey matter volume, cortical thickness or surface area. Using these approaches, it is possible to monitor the thinning of grey matter, which likely reflects the death of neuronal cell bodies at advanced stages of neurodegeneration. Synaptic disruption is an early event in Alzheimer’s disease (Sperling *et al*., 2011), and functional networks may have the ability to compensate the impact of neurodegeneration on cognitive symptoms (Franzmeier *et al*., 2017). For these reasons, intrinsic functional connectivity from resting-state fMRI is an emerging Alzheimer’s disease biomarker that holds promise for early diagnosis (Sperling *et al*., 2011; Badhwar *et al*., 2017). To analyze resting-state fMRI, select regions in canonical brain networks previously established in the literature are generally considered (1b). An individual resting-state fMRI connectivity map can be generated for different networks, with the default-mode, limbic, and salience networks being the key components affected by Alzheimer’s disease (Badhwar *et al*., 2017) (1b). Structural and functional brain maps enter a subtyping procedure, which identifies groups of individuals with homogeneous brain maps (1c).The number of subtypes are defined a priori or through various metrics for model selection (Seghier, 2018) (for example N=3 in 1c). A subtype map is generated by averaging the maps within each subgroup and subtracting the grand average (i.e. demeaned) to emphasize the features of the subtype. Chi square statistics are applied to identify groups that include a greater number of Alzheimer’s disease patients than expected by chance (illustrated by a “* AD” annotation for subtype 2 in 1c). In 1d, the subtyping procedure was applied on maps of grey matter density from CN and Alzheimer’s disease dementia individuals in the ADNI database (N=377). Four out of seven subtypes were identified as Alzheimer’s disease dementia-related (results adapted from Tam and colleagues (Tam *et al*., 2018)). Three subtypes were consistent with previous reports: posterior (or temporo-occipito-parietal-predominant), diffuse, and temporal (or medial temporal-predominant) atrophy subtypes. A novel language atrophy subtype was also identified. In 1e, the subtype procedure was applied to resting-state fMRI data collected on CN, MCI, and Alzheimer’s disease dementia individuals in a dataset pooling ADNI2 with several independent samples (N=130). Three subtypes were extracted for three resting-state networks known to be impacted by Alzheimer’s disease: default-mode, salience, and limbic. One Alzheimer’s disease dementia/MCI-related subtype was found for each network. The salience and default-mode followed similar patterns: increased within-network connectivity, and a lower (negative) connectivity between networks. The limbic subtypes showed lower connectivity with frontal regions, and increased connectivity with occipital regions. Results adapted from Orban et al. (Orban *et al*., 2017). The section on “Brain Subtypes” compares results from the above-mentioned and other studies with similar approaches and objectives. Supplementary Table S1 provides detailed characteristics of the 12 neuroimaging subtyping studies (structural MRI and resting-state fMRI) that met our search criteria. Abbreviations: BOLD (blood oxygen level dependant sigal)

**Figure 2:**
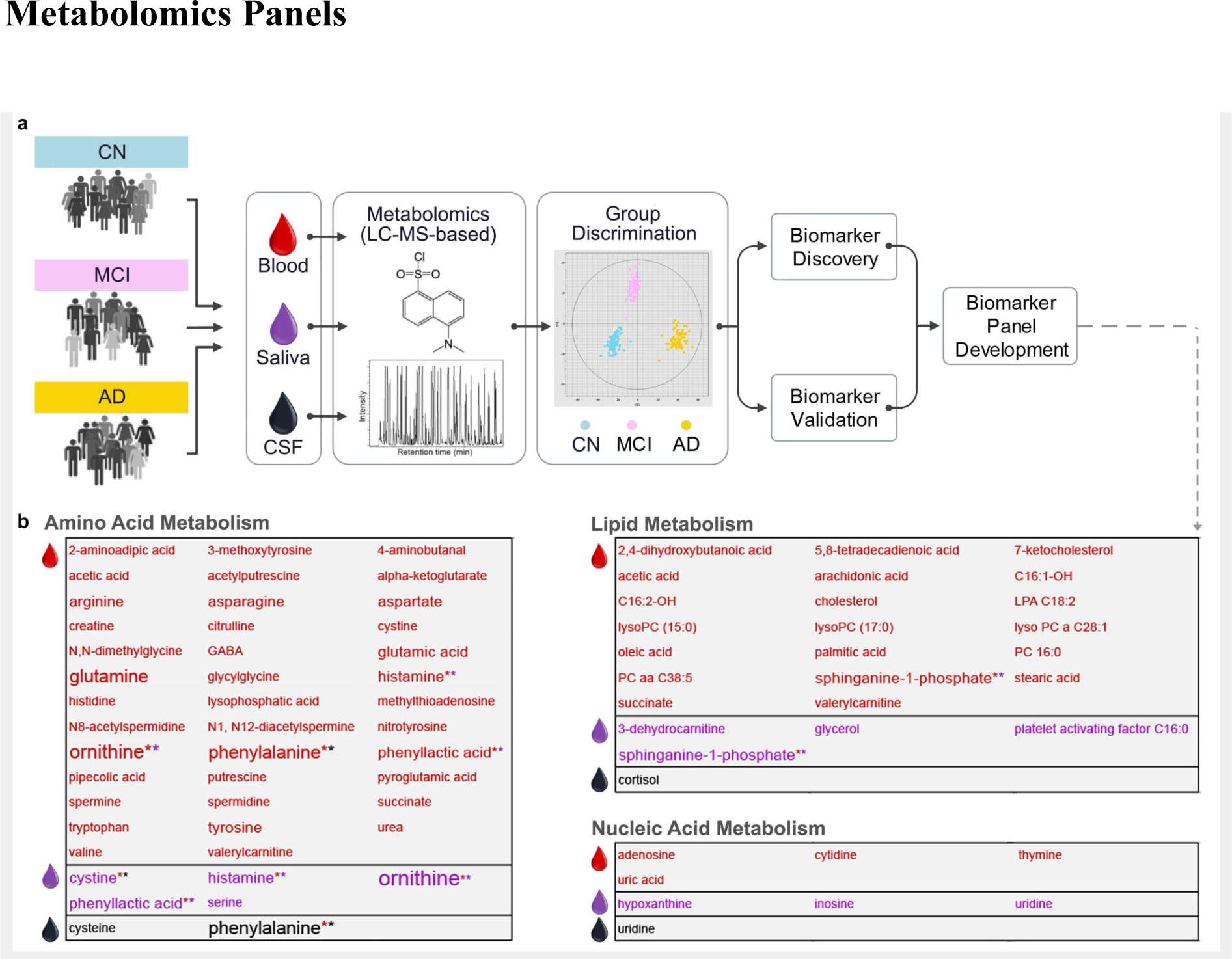
A typical Alzheimer’s disease metabolomics biomarker discover pipeline. Metabolomics is a relatively recent addition to the systems biology toolkit for the study of NDDs of aging (Wilkins and Trushina, 2017). It encompasses the global study of small-molecules (50-1500 daltons in mass), that are substrates and products of metabolism. Together, these metabolites (e.g. amino acids, antioxidants, vitamins) represent the overall physiological status of the organism. An individual’s metabolic activity is influenced by an individual’s genotype and environment (Kaddurah-Daouk *et al*., 2011). Analysis of the metabolome, therefore, provides an opportunity to study the dynamic molecular phenotype of an individual. Untargeted metabolomics approaches are increasingly used to compare two or more groups (e.g. Alzheimer’s disease dementia and CN participants) and identify metabolite profiles associated with a disease. These profiles provide insight into underlying disease mechanisms, as well as constitute candidates for biomarker discovery and drug development. In the field of Alzheimer’s disease research, metabolomics studies (targeted and untargeted) over the last decade have examined several biofluids and tissues, including serum, plasma, CSF, saliva, urine, and brain tissue (Wilkins and Trushina, 2017). Technologies include NMR (nuclear magnetic resonance) spectroscopy and mass spectrometry. In 2a, a typical Alzheimer’s disease metabolomics biomarker discovery pipeline using MS-LC (mass spectrometry-liquid chromatography) is depicted. Subsequent to metabolite extraction, identification, and quantification, most studies apply multivariate statistical methods to the metabolome data to identify the top discriminant metabolites. These can be further combined into metabolite-panels to increase discriminative power (i.e. sensitivity and specificity) in Alzheimer’s disease prediction and progression (Liang *et al*., 2015, 2016; Huan *et al*., 2018). Significant discriminative power is commonly tested with the Receiver Operating Characteristic curve analysis (area under the curve or AUC values). Discriminant metabolite-panels are then validated in independent samples. Following discriminant metabolite(s) discovery, researchers conduct pathway and network analyses, which provide crucial mechanistic insights into the sequences of processes leading to the heterogeneous phenotypes of neurodegeneration. Pathway analysis focus on identifying sequences of processes that lead to the presence of a discriminant metabolite. Network analysis examine how discriminant metabolites are connected to each other within Alzheimer’s disease and related dementias. In 2b, we show the three main metabolic pathways (namely, amino acid, lipid and nucleic acid) that 90 Alzheimer’s disease-associated metabolites in our review (*N*=11 publications, Supplementary Table S2) were found to belong. The text color indicates the biofluid metabolome each metabolite was identified in: red = serum or plasma, purple = saliva, black = CSF. A larger font size indicates that the metabolite was identified in more than one study (see Supplementary Table S3 for details) The maximum number of studies a metabolite was detected in our review was four. * indicates that the metabolite was identified in more than one biofluid:* * indicates presence in plasma or serum and saliva, * * indicates presence in plasma or serum and CSF. Abbreviations: AD, Alzheimer’s disease; CN, cognitively normal; MCI, mild cognitive impairment.

### Genomics

Search term combinations used for the genomics were: (Polygenic Risk Scale OR Polygenic Risk Index OR Genetic Risk Score OR Genetic Risk Scale OR Genetic Risk Index) AND (Alzheimer Disease OR Alzheimers Disease OR Alzheimer’s Disease), which identified 264 potential papers. To these we applied the “common” exclusion criteria, followed by exclusion of single-gene studies. In total 17 studies were reviewed. References used for the compilation of the genes in Supplementary Table S4 are provided in the Supplemental Reference section.

### Anatomical subtypes

The spatial distribution of brain atrophy on structural MRI is highly heterogeneous in MCI, and Alzheimer’s disease dementia patients (Nettiksimmons *et al*., 2014; Poulakis *et al*., 2018). Using data-driven clustering algorithms, 11 studies have attempted to subtype and characterize this inherent heterogeneity (Supplementary Table S1). Seven studies reported at least three distinct atrophy subtypes in Alzheimer’s disease dementia (Noh *et al*., 2014; Hwang *et al*., 2016; Park *et al*., 2017; Poulakis *et al*., 2018; ten Kate *et al*., 2018), or mixed (Alzheimer’s disease dementia and CN) cohorts (Varol *et al*., 2017; Tam *et al*., 2018). Subtypes were generally consistent across studies, and can be described as diffuse, medial temporal-predominant (temporal), and temporo-occipito-parietal-predominant (posterior) (Fig. 1d). They were generated by applying (1) hierarchical agglomerative clustering using Ward’s clustering linkage (Noh *et al*., 2014; Hwang *et al*., 2016; Tam *et al*., 2018), Louvain clustering (Park *et al*., 2017), random forest clustering (Poulakis *et al*., 2018), or non-negative matrix factorization (ten Kate *et al*., 2018) on cortical thickness or grey matter density maps, or (2) clustering on grey matter density maps using a novel approach called HYDRA (Varol *et al*., 2017). Good agreement across studies may, in part, reflect usage of the same data sample (ADNI) for subtype identification in four studies (Hwang *et al*., 2016; Varol *et al*., 2017; Poulakis *et al*., 2018; Tam *et al*., 2018). Some studies report two-subtype decomposition (Dong *et al*., 2016; Malpas, 2016), but these lack inter-study consensus. Using model-based clustering on regional cortical thickness measures from ADNI, Malpas reported normal and atrophic-entorhinal subtypes in a sample including Alzheimer’s disease dementia, and CN individuals (Malpas, 2016). The atrophic-entorhinal subtype demonstrated considerable heterogeneity in entorhinal thickness, suggesting the presence of additional subtypes (Malpas, 2016). Dong et al. reported limbic-insular, and parietal-occipital atrophy subtypes using CHIMERA clustering on brain volume data from Alzheimer’s disease dementia and CN ADNI participants (Dong *et al*., 2016). In a separate study, the same group reported four atrophy subtypes using CHIMERA: normal, temporal, and two diffuse subtypes - one with predominant temporal involvement (diffuse-temporal), and one without (diffuse) (Dong *et al*., 2017). Visually, the diffuse subtype from CHIMERA shared some overlap with the posterior subtype described previously. In general, the reported subtypes (Dong *et al*., 2017) fit better with the three subtype solution, considering that, unlike previous studies, the CHIMERA study included CN individuals. Finally, Tam et al. identified a fourth atrophy subtype involving several language-related areas (Tam *et al*., 2018).

The choice of the number of subtypes is to some degree arbitrary. Two studies showed that their three subtypes could be decomposed into six (Noh *et al*., 2014), or more (Tam *et al*., 2018) homogeneous groups. Finally, an additional study looking at heterogeneity with a linear mixture model, instead of a discrete cluster analysis, showed that most individuals tend to express varying levels of multiple subtypes (Zhang *et al*., 2016). Continuous measures of subtype similarity are thus more advisable than discrete assignment (Zhang *et al*., 2016; Tam *et al*., 2018).

We now highlight various associations between Alzheimer’s disease markers/risk-factors and the three atrophy subtypes consistently reported. In three studies, Alzheimer’s disease dementia patients with the posterior subtype were reported to be the youngest, and had the earliest age-at-onset (Noh *et al*., 2014; Hwang *et al*., 2016; Park *et al*., 2017). They also demonstrated greater PET-detectable amyloidosis (Hwang *et al*., 2016), and pathological levels of CSF Aβ42 and tau (Noh *et al*., 2014; Varol *et al*., 2017; ten Kate *et al*., 2018). Differences across subtypes were reported with FDG-PET-detectable glucose hypometabolism (Hwang *et al*., 2016), and white matter hyperintensities (ten Kate *et al*., 2018). Subtype-specific associations with Alzheimer’s disease-related genes were observed, specifically, *APOE* (Noh et al., 2014; Varol et al., 2017), *CD2AP (CD2-associated protein)* (Varol et al., 2017), *SPON1 (Spondin-1)* (Varol et al., 2017), *LOC390956* or *PPIAP59 (peptidyl-prolyl cis-trans isomerase A pseudogene)* (Varol et al., 2017), though the association with *APOE* was not consistently found (Hwang *et al*., 2016). Associations of subtypes with cognition were observed for global (Varol *et al*., 2017; Tam *et al*., 2018) and domain-specific (e.g. episodic memory) (Noh *et al*., 2014; Park *et al*., 2017; Poulakis *et al*., 2018; ten Kate *et al*., 2018) measures, but not by all studies (Hwang *et al*., 2016). Associations between subtypes and sex were found to be significant in two (Noh *et al*., 2014; Varol *et al*., 2017) of four (Noh *et al*., 2014; Hwang *et al*., 2016; Varol *et al*., 2017; Tam *et al*., 2018) studies.

### Functional subtypes

By coupling cluster analysis and resting-state functional MRI, a preprint report by Orban et al. (Orban *et al*., 2017) investigated connectivity subtypes in CN, MCI, and Alzheimer’s disease dementia patients (Supplementary Table S1). They noted associations between functional connectivity subtypes and cognitive symptoms in the default-mode, limbic, and salience networks in MCI, and Alzheimer’s disease dementia patients (Fig. 1e). Limbic subtypes were also associated with Alzheimer’s disease biomarkers (CSF Aß42 levels, *APOE4* genotype) in an independent cohort at increased risk for familial Alzheimer’s disease, suggesting that functional connectivity subtypes may be sensitive to the presence and progression of preclinical disease (Orban *et al*., 2017).

### Summary

Our review found convergent evidence of distinct brain atrophy subtypes in Alzheimer’s disease dementia patients, including at least three data-driven Alzheimer’s disease atrophy subtypes: diffuse, temporal, and posterior. These structural subtypes seem to associate with established biomarkers, risk factors, and clinical symptoms of Alzheimer’s disease, as well as cognitive subtypes: temporal subtype with memory impairment, and diffuse subtype with impaired executive function (Zhang *et al*., 2016). The picture emerging from fMRI data is one of aberrant between-network connectivity initiating in the mesolimbic network at the preclinical stage and propagating to the salience and default-mode network with Alzheimer’s disease progression (Orban *et al*., 2017).

### Metabolite Panels

We reviewed six Alzheimer’s disease studies that constructed metabolite panels from top discriminant metabolites in biofluids: 2 plasma, 1 serum, 2 saliva, and 1 CSF (Supplementary Table S2). Using plasma metabolome data, Wang et al. (Wang *et al*., 2014) constructed a six-metabolite panel to discriminate Alzheimer’s disease dementia from CN, and a five-metabolite panel to discriminate amnestic MCI from CN. Arachidonic acid, N,N-dimethylglycine, and thymine were present in both panels. Association of panel metabolites with lipid, amino acid, or nucleic acid metabolism suggested specific metabolic deregulations in Alzheimer’s disease. Mapstone et al. (Mapstone *et al*., 2017) used a 12-plasma-metabolite panel to discriminate the following cohorts from CN: older adults with superior memory; amnestic MCI + Alzheimer’s disease dementia patients; and participants who converted to amnestic MCI or Alzheimer’s disease dementia in approximately two years. Similar to Wang et al. (Wang *et al*., 2014), several panel metabolites were associated with lipid or amino acid metabolism. Panel metabolites were also found to be constituents of pathways regulating oxidative stress, inflammation, and nitric oxide bioavailability. Using serum metabolome data, Liang et al. (Liang *et al*., 2016) identified a panel of two lipid metabolites (spinganine-1-phosphate, 7-ketocholesterol) that discriminated MCI from Alzheimer’s disease dementia. Sphinganine-1-phosphate was also present in a three-metabolite panel constructed from the salivary metabolome, and discriminated Alzheimer’s disease dementia patients from CN (Liang *et al*., 2015). The other two panel members, ornithine and phenyllactic acid, were amino acid metabolites (Ogata *et al*., 1999) with links to the same oxidative stress pathway reported by Mapstone et al. (Mapstone *et al*., 2017) A separate study using saliva reported a seven-metabolite panel that discriminated pre-dementia (i.e. five years prior to dementia onset) from CN (Figueira *et al*., 2016). Metabolites were associated with amino acid, lipid, or energy metabolism. Czech et al. (Czech *et al*., 2012) assessed multiple combinations of 16 CSF metabolites to discriminate Alzheimer’s disease dementia from CN. Highest discrimination was obtained with a 5-metabolite panel consisting of cortisol and amino acids. Our focused review of Alzheimer’s disease-associated metabolite panels highlight that the majority of discriminant molecules detected in biofluids are involved in amino acid, lipid, or nucleic acid metabolism (Fig. 2b).

### Metabolomics Pathways and Networks

We reviewed five Alzheimer’s disease studies that followed up non-targeted metabolomics research in biofluids with pathway or network analyses: 2 plasma, 1 plasma plus CSF, and 2 serum (Supplementary Table S2). In plasma, de Leeuw et al. (de Leeuw *et al*., 2017) identified 26 metabolites comprised of mainly amino acids and lipids with significantly altered levels in Alzheimer’s disease dementia patients. Network analyses suggested a shift in Alzheimer’s disease towards amine and oxidative stress compounds, known to cause imbalances in neurotransmitter production, Aβ generation, and neurovascular health. Perturbations in amino acid metabolism (interlinked polyamine and L-arginine pathways) was also demonstrated in the plasma metabolome of MCI to Alzheimer’s disease dementia converters (Graham *et al*., 2015). Changes in polyamine and L-arginine metabolism have been linked to neurotoxicity, and deregulations in genesis and/or death of neural cells and neurotransmitter production (Graham *et al*., 2015). Other metabolic pathways notably impacted were cholesterol, glucose, and prostaglandin (Graham *et al*., 2015). Cholesterol metabolism (specifically cholesterol and sphingolipids transport) was also found to be abnormal in both plasma and CSF from Alzheimer’s disease dementia patients (Trushina *et al*., 2013). In serum, metabolism of amino acids dominated the top pathways altered in Alzheimer’s disease dementia patients in one study (González-Domínguez *et al*., 2015), a finding in line with plasma metabolome data (de Leeuw *et al*., 2017). A second study in serum reported a 3-metabolite panel predictive of progression from MCI to Alzheimer’s disease dementia (within 27±18 months), with major contribution from up-regulated 2,4-dihydroxybutanoic acid, a metabolite potentially overproduced during hypoperfusion-related hypoxia (Orešič *et al*., 2011). Upregulation of the pentose phosphate pathway in progressors further supported the involvement of secondary hypoxia in Alzheimer’s disease pathogenesis. More glucose is metabolized via the pentose phosphate pathway in the brain under hypoxic conditions.

### Summary

Overall, metabolite panels, and metabolomics pathway and network analyses provide the following insights: (1) discriminant Alzheimer’s disease-associated metabolites may be narrowly or broadly interconnected (Wilkins and Trushina, 2017); (2) metabolomes of different biofluids provide convergent and biofluid-related mechanistic insights into Alzheimer’s disease pathology (Trushina *et al*., 2013); (3) genotype-associated (e.g. *APOE* status) differences in preclinical and clinical groups suggest different routes to Alzheimer’s disease (de Leeuw *et al*., 2017), (4) neurodegenerative disease subtypes can be characterized by metabolomics analyses (de Leeuw *et al*., 2017).

### Polygenic Risk Score Approach or PRS

Constructed of multiple SNPs that implicate one or more biological mechanisms, PRS (Fig. 3) are better at discriminating Alzheimer’s disease from CN than single-gene analysis (Escott-Price *et al*., 2017; Torkamani *et al*., 2018). We reviewed 11 Alzheimer’s disease PRS studies (Supplementary Table S4, S5), the majority comprised of GWAS-detected SNPs. To identify genetic risk beyond that of *APOE* alone, several studies assessed PRS with (*APOE*-PRS) and without (non-*APOE*-PRS) *APOE*. Desikan et al. (Desikan *et al*., 2017) found that an *APOE*-PRS associated with age-at-onset of Alzheimer’s disease symptoms, decreased Aβ and increased tau in CSF, and increased atrophy, tau, and Aβ load in brain. *APOE*-PRS also associated with plasma inflammatory markers in Alzheimer’s disease patients (Morgan *et al*., 2017). An *APOE*-PRS including a rare *TREM2 (triggering receptor expressed on myeloid cells 2)* variant discriminated Alzheimer’s disease dementia and CN, with increasing scores associating with decreasing age-at-onset, and CSF Aβ42 (Sleegers *et al*., 2015). Discriminative power of *APOE*-PRS was found to improve with diagnostic accuracy, as demonstrated using a pathologically confirmed Alzheimer’s disease cohort (Escott-Price V, Myers, AJ, Huentelman M, Hardy, J, 2017). In four separate studies, a non-*APOE-*PRS was reported to discriminate between Alzheimer’s disease dementia and CN (Xiao *et al*., 2015), as well as associate with MCI (Adams *et al*., 2015), increased risk of Alzheimer’s disease dementia (Adams *et al*., 2015; Chouraki *et al*., 2016; Tosto *et al*., 2017), and earlier Alzheimer’s disease onset (Tosto *et al*., 2017). Inclusion of *APOE* either resulted in a modest increase in discriminative power (Xiao *et al*., 2015), stronger clinical or biomarker associations (Adams *et al*., 2015; Chouraki *et al*., 2016), or had no additional effect (Tosto *et al*., 2017). In another non-*APOE*-PRS study in Alzheimer’s disease patients, PRS scores correlated negatively with CSF Aβ42 levels, and positively with temporal cortex Aβ pathology, and γ-secretase activity (Martiskainen *et al*., 2015). Naj et al. (Naj *et al*., 2014) found that though *APOE* contributed to 3.7% of age-at-onset variability in Alzheimer’s disease dementia patients, adding a non-*APOE-*PRS accounted for an additional 2.2%. Overall, Alzheimer’s disease heritability has a large polygenic contribution beyond *APOE*, which makes PRS approaches pivotal for Alzheimer’s disease-risk prediction (Escott-Price *et al*., 2015).

**Figure 3:**
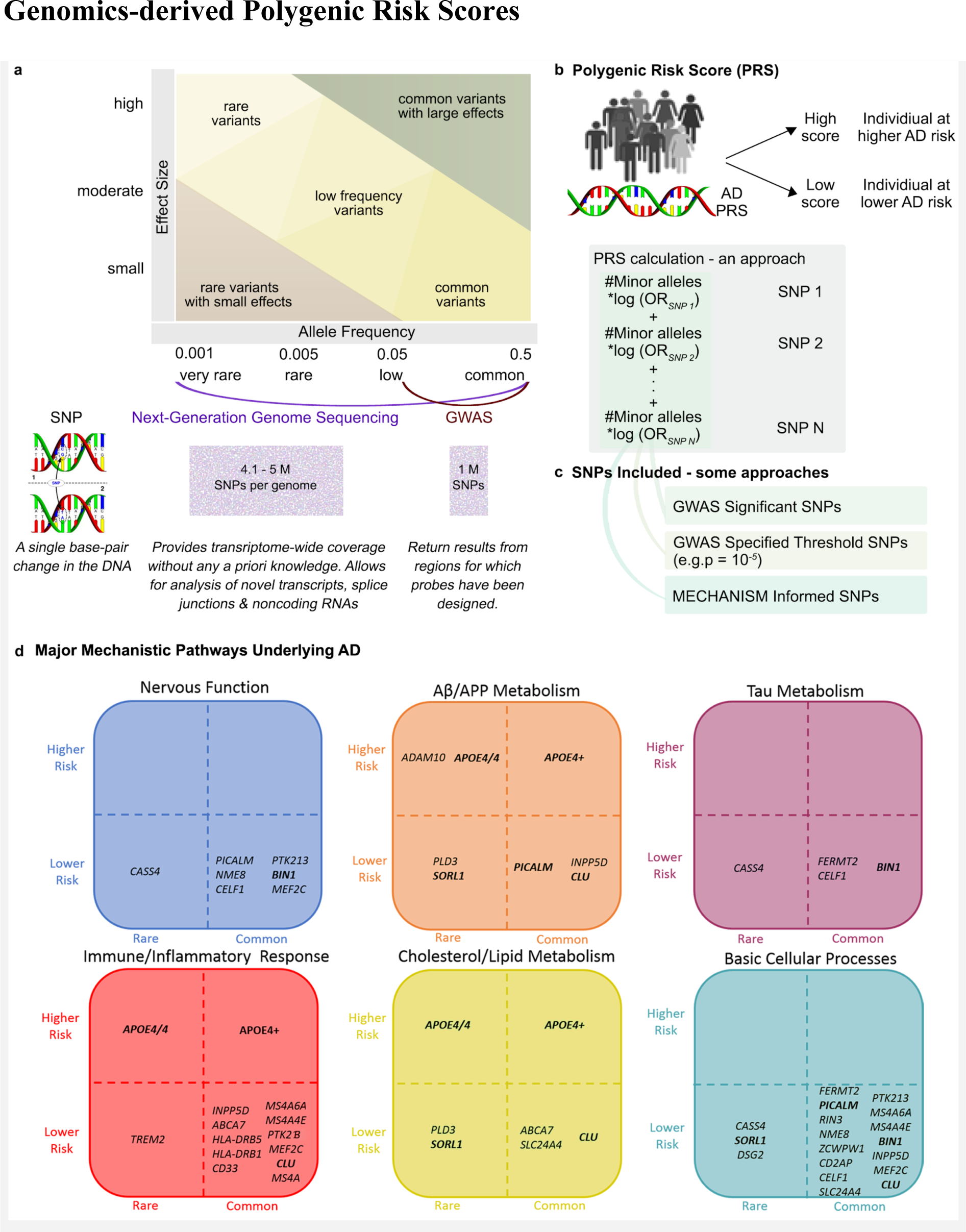
Polygenic Risk Scores. High-throughput genotyping technologies have revolutionized studies in diseases with complex genetics by enabling detection of common genetic variants with low effect sizes, and rarer variants with relatively higher effect sizes (see 3a). In Alzheimer’s disease, the prevalent late-onset variant is genetically complex and demonstrates high heritability (up to 80%) (Gatz *et al*., 2006), whereas the early-onset familial variant is deterministically driven by single gene mutation(s) in *PSEN1* (presenilin 1), *PSEN2* (presenilin 2) or the *APP* (amyloid precursor protein) (Guerreiro *et al*., 2013). The genetics of late-onset Alzheimer’s disease has been predominantly investigated using GWAS. Designed to rapidly scan for statistical links between a set of known SNPs and a phenotype of interest, GWAS can identify common variants with MAF greater than 5% (Torkamani *et al*., 2018) (see 3a). Up to 24 key Alzheimer’s disease-risk genes have been identified using GWAS (Supplementary Table S4). Identification of rarer Alzheimer’s disease-associated SNPs (MAF >0.5% and <5%), that often escape detection with GWAS, is being enabled by NGS technologies, such as whole-exome sequencing and targeted resequencing of disease-associated genes (Bras *et al*., 2012; Masellis *et al*., 2013) (see Supplementary Table S4 for examples). NGS technologies provide transcriptome-wide coverage without requiring any a priori knowledge of SNPs (3a). To date, Alzheimer’s disease prediction using individual high-throughput genotyping technologies identified risk genes have been predominantly non-significant, with the exception of *APOE*, which accounts for up to 30% of the genetic risk (Daw *et al*., 2000). Therefore, the search for risk genes beyond *APOE* now include polygenic risk score (PRS, also referred to as genetic risk scores, risk indexes or scales) approaches (3b). A PRS is a calculation (e.g. weighted sum) based on the number of risk alleles carried by an individual, where the risk alleles and their weights are defined by GWAS-detected loci and their measured effects (Torkamani *et al*., 2018). In the most common scenario, only SNPs reaching conventional GWAS significance (*p* < 5 ⨯ 10^−8^) are included (3c). A threshold lower than the genome-wide statistical significance (e.g. *p* = 10^−5^) can also be used to improve or estimate total predictability (Torkamani *et al*., 2018) (3c). SNPs representing multiple hits among Alzheimer’s disease risk genes from one or more major mechanistic pathways can also be included into a PRS (3c). Displayed are six main mechanistic clusters, each populated by genetic variants thought to represent the cluster (3d). Genetic variants have been placed within the cluster according to population frequency (horizontal axis) and level of estimated risk (vertical axis). For example, an Aβ/APP metabolism cluster is made up of rare *ADAM10 (a disintegrin and metalloproteinase domain-containing protein 10)* and common *APOE4+*)higher risk genes, and rare *PLD3 (phospholipase D family member 3)* and common *PICALM (phosphatidylinositol binding clathrin assembly protein)* lower risk genes. Some genes are involved with multiple mechanisms as can be seen for *PICALM*’s involvement in nervous function, basic cellular processes, and Aβ/APP metabolism. As implied in the figure, when creating PRS, it may be very useful to select genes within mechanistic groups, and select groups depending on the purpose of the research. In sum, PRS reflect a large number of SNPs and a complex set of biological mechanisms related to Alzheimer’s disease pathogenesis, and can improve the precision of early Alzheimer’s disease risk or diagnosis (Desikan *et al*., 2017; Escott-Price *et al*., 2017; Morgan *et al*., 2017).

### Mechanism-based Interaction and Network Approaches

Alzheimer’s disease-risk genes can be clustered into functional/mechanistic pathways (Fig. 3d), and the information gained utilised to improve Alzheimer’s disease discrimination and/or risk prediction (Gaiteri *et al*., 2016; Hu *et al*., 2017). We reviewed six mechanism-based Alzheimer’s disease studies (Supplementary Table S5). Functional variants of Alzheimer’s disease GWAS-significant SNPs (e.g. *CELF1* or *CUGBP Elav-like family member 1*) was reported to associate with human brain expression quantitative trait loci, and preferentially expressed in specific cell-types (e.g. microglia) (Karch *et al*., 2016). Rosenthal et al. (Rosenthal *et al*., 2014) highlighted the potential regulatory functions of non-coding Alzheimer’s disease GWAS SNPs. Protein-protein interaction network analyses highlighted that Alzheimer’s disease-risk genes whose protein-products interact physically may be under positive evolutionary selection (e.g. *PICALM* or *phosphatidylinositol binding clathrin assembly protein, BIN1* or *bridging integrator 1, CD2AP* or *CD2 associated protein, EPHA1* or *EPH receptor A1*) (Raj *et al*., 2012). Ebbert et al. (Ebbert *et al*., 2014) reported that while an *APOE*-PRS did not improve discrimination of Alzheimer’s disease from CN over *APOE*, a model allowing for epistatic interactions between SNPs increased discriminative power. Patel et al. (Patel *et al*., 2016) applied a stratified false discovery rate approach, used to increase GWAS power by adjusting significance levels to the amount of overall signal present in data, to identify gene networks and provide links with sMRI phenotypes: e.g. linking genes involved in transport (e.g. *SLC4A10* or *solute carrier family 4 member 10, KCNH7* or *potassium voltage-gated channel subfamily H member 7*) with hippocampal volume. Huang et al. (Huang *et al*., 2018) integrated Alzheimer’s disease GWAS genes with human brain-specific gene network using machine learning to identify additional Alzheimer’s disease-risk genes.

### Summary

In sum, PRSs may contribute substantially to accounting for the genetic variability that distinguishes Alzheimer’s disease from MCI and CN groups. They may also be used to probe genetic underpinnings of Alzheimer’s disease subtypes as well as related and disparate NDDs. Thus far, research reporting PRSs in relation to conversion rates of CN or MCI to Alzheimer’s disease dementia have been mixed (Adams *et al*., 2015; Lacour *et al*., 2017), but early PRS prediction of cognitive trajectories and clinical outcomes have also been reported (Sapkota and Dixon, 2018).

## Discussion

### Roadmap for assembling multiomics measures into diagnostic tool

#### On the complementarity of multiomics biomarkers

Individuals clinically diagnosed with an NDD of aging (e.g. Alzheimer’s disease) exhibit varying loads of neurodegenerative markers (e.g. Aβ, tau, alpha-synuclein, brain atrophy, vascular abnormalities) (Beach *et al*., 2012; Robinson *et al*., 2018). Single-domain omics biomarkers can, to an extent, characterize this heterogeneity *in vivo*. Some data-driven brain atrophy subtypes parallel established clinical diagnoses. For example, the posterior atrophy subtype is evocative of the PCA Alzheimer’s disease variant, and the language atrophy subtype of the lvPPA variant (Ossenkoppele *et al*., 2015). An active area of research is to determine to what degree the “bottom-up”, fully automated and data-driven subtypes match with established “top-down” clinical assessments, which usually start with cognitive symptoms and then incorporate specific neuroimaging characteristics, such as left temporoparietal atrophy in lvPPA (Ossenkoppele *et al*., 2015). The fact that reviewed studies included participants with typical late-onset Alzheimer’s disease dementia, and not atypical variants such as PCA, suggest that specific brain atrophy phenotypes comprise a spectrum of involvement that may overlap a clinical label, but not associate uniquely with one. It is unclear how functional connectivity subtypes tie in with atrophy subtypes, although they both associate with clinical diagnoses and biomarkers and risk factors of Alzheimer’s disease (Zhang *et al*., 2016; Orban *et al*., 2017). Although the propagation of functional dysconnectivity parallels the Braak staging of Alzheimer’s disease, the mix of connectivity increases and decreases observed in patients may reflect transient compensatory mechanisms as well as neurodegeneration (Badhwar *et al*., 2017). To date, we are unaware of studies that have associated data-driven Alzheimer’s disease brain subtypes with PRS and/or metabolite panels, but our review strongly supports such coordinated multiomics approaches. For example, an Alzheimer’s disease PRS comprised of genes linked to lipid metabolism and inflammatory response may associate with panels comprised of metabolites involved in corresponding pathways. One could further speculate whether the resulting inflammation may cause the diffuse atrophy subtype, and trigger specific functional compensatory mechanisms. Testing these hypotheses will require a cohort that is both deeply phenotyped and captures the entire spectrum of age-related dementia.

#### A roadmap for parsing heterogeneity in neurodegeneration

A data-driven characterization of heterogeneity across the NDDs of aging will require cohorts representative of the spectrum of neurodegeneration. The cohort assembled by the Canadian Consortium on Neurodegeneration in Aging (CCNA, http://ccna-ccnv.ca/) provides a new opportunity to study the full spectrum of age-related dementia. By 2019, the cohort will include 2,310 individuals (ages 50-90) featuring the following cognitive conditions: Alzheimer’s disease, vascular, Lewy Body, Parkinson’s, frontotemporal, and mixed etiology dementias, as well as subjective cognitive impairment, MCI, vascular MCI, and CN (Fig. 4).

**Figure 4:**
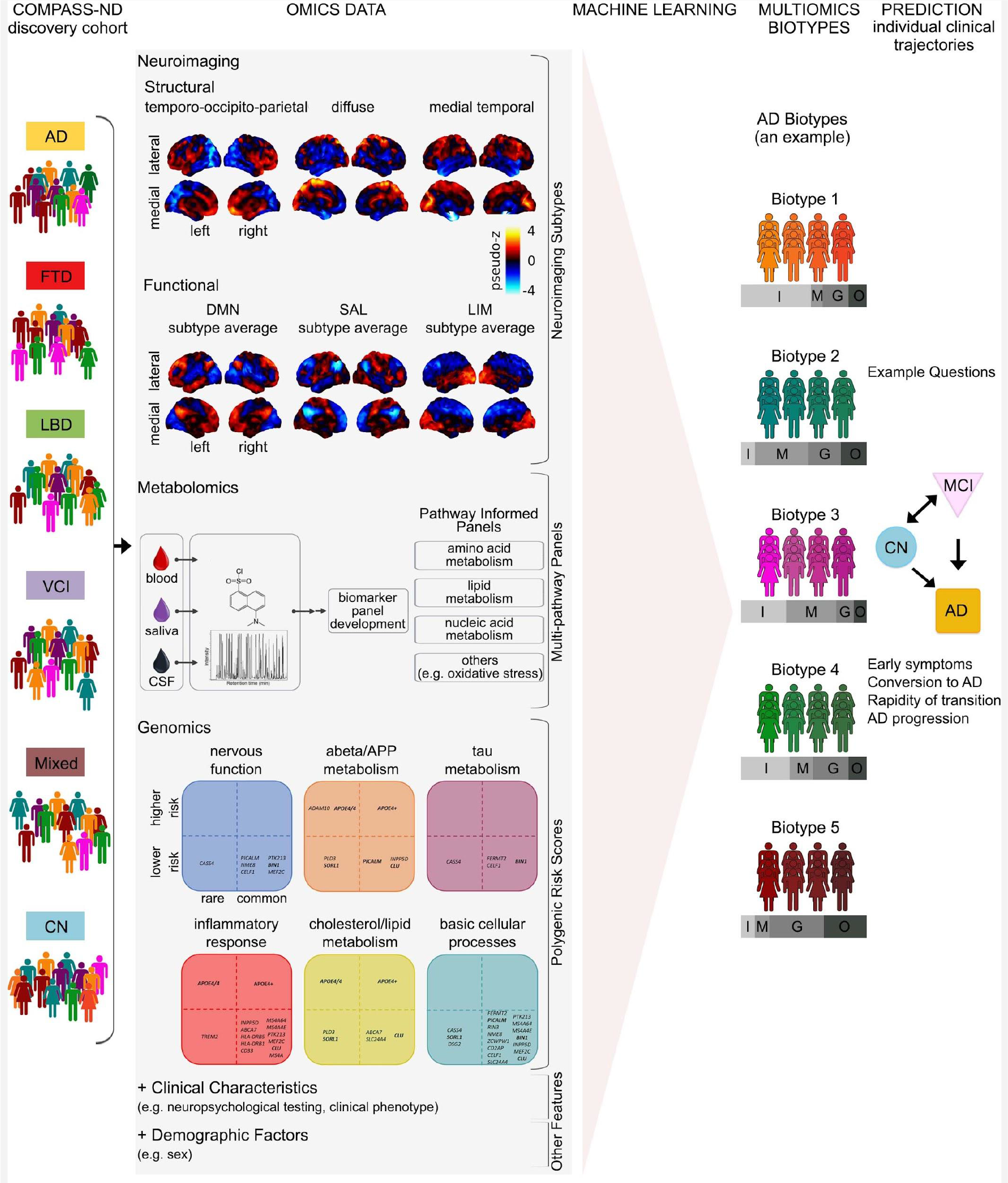
proposed roadmap to discovering multiomics Alzheimer’s disease biomarkers. *Panel 1 (COMPASS-ND):* The COMPASS-ND cohort is comprised of people with various types of dementia or cognitive complaints, as well as healthy, cognitively normal individuals. *Panel 2 (Omics data):* Performing dimension reduction for omics data. Featured as examples are some of the results of our review of the Alzheimer’s disease literature as presented earlier in the paper. *Panels 3-5 (Machine Learning, Multiomics Biotypes and Prediction):* demonstrate how signatures of neurodegeneration derived from the integration of multiomics data using machine learning techniques will better identify individuals on an Alzheimer’s disease spectrum trajectory. While our proposed roadmap addresses multiomics biomarkers for Alzheimer’s disease, a similar approach can be used for other neurodegenerative diseases of aging. Abbreviations: Alzheimer’s Disease (AD), FrontoTemporal Dementia (FTD), Lewy Body Disease (LBD), Vascular Cognitive Impairment (VCI), Mixed etiology dementia (Mixed), Cognitively normal (CN), Subjective Cognitive Impairment (SCI), Mild Cognitive Impairment (MCI), Imaging features (I), Metabolic features (M), Genomics features (G), clinical and demographic features (O).

The cohort composition ensures that age-related dementias are more or less equally represented, even for less prevalent dementia types (e.g. frontotemporal dementia). Participants will be deeply phenotyped with extensive clinical, neuropsychological, neuroimaging, biospecimen, and neuropathological assessments.

In Fig. 4 we present a roadmap for a multiomics approach to heterogeneity in NDD. We begin with a heterogeneous clinical cohort design that enables the discovery of subgroups sharing a common signature across multiple omics domains (biotypes) that are highly predictive of the clinical status and evolution of individual patients. Multiomics biotypes will be complemented by other important variables such as sex, presence of Aβ and tau deposits, and vascular abnormalities. Machine learning tools will be applied to identify an optimal combination of different biotypes and explanatory variables that either discriminate different clinical cohorts, or are predictive of future progression of specific symptoms (Fig. 4).

#### Towards highly predictive multiomics signatures

Biomarkers of Alzheimer’s disease dementia demonstrate limited predictive power in the prodromal phase (Rathore *et al*., 2017). For example, Korolev et al. (Korolev *et al*., 2016) reached about 80% accuracy (specificity 76%, sensitivity 83%) by including cognitive, multimodal imaging, and plasma-proteomics measures in a predictive model of progression. The best models include a combination of cognitive, sMRI, FDG-PET, and/or amyloid-PET measures (Rathore *et al*., 2017). A substantial proportion of patients identified as progressors, even by the best model, will remain stable over time. Given a 30% baseline rate of progressors in an MCI cohort, and a prediction achieving a sensitivity and specificity of 80%, 37% of patients identified as progressors by the model will remain stable. This limited positive predictive value is likely explained by sample heterogeneity. For example, Dong et al. (Dong *et al*., 2017) reported two atrophy subtypes with higher-than-expected rates of progression to dementia, but one subtype was much more at risk than the other. Thus, an important first step to precision medicine is to identify specific subsets of patients where an accurate prediction can be made.

Multiomics signatures can be used to improve the accuracy of early prognosis, but they also capture a range of information, ranging from brain networks targeted by the disease, metabolic abnormalities in specific pathways, and distinct genetic backgrounds. The multiomics signature thus may also help elucidate the specific pathophysiological pathways involved. Overall, multiomics biomarkers have the potential to reshape clinical diagnosis, and define new “bottom up” cohorts based on markers of underlying pathologies to design and evaluate drugs.

## Data Availability

Not applicable

## Abbreviations

Aβ or Aβ42: amyloid beta or amyloid beta 42
APOE: apolipoprotein E
ADNI: Alzheimer’s Disease Neuroimaging Initiative
CN: cognitively normal
FDG: fluorodeoxyglucose
fMRI: functional MRI
GWAS: genome-wide association studies
lvPPA: logopenic progressive aphasia
MAF: minor allele frequency
MCI: mild cognitive impairment
NDDs: neurodegenerative diseases
NGS: next-generation genome sequencing
PCA: posterior cortical atrophy
PET: positron emission tomography
PRS: polygenic risk score
SNPs: single nucleotide polymorphisms

## Author Contributions

AB, PB, RAD, GPM, and SS designed and wrote the manuscript. AB, GPM and SS performed literature search and compiled tables. AB, PB, RAD, GPM and SS made the figures and MM provided input. MM, LL, SD, HC, and SEB helped with the writing and interpretation of specific sections of the manuscript.

## Acknowledgements

We would like to thank Dr. Sridar Narayanan for helpful comments on the manuscript.

## Supplementary Material

We have provided 5 tables in supplementary material: Supplementary Tables S1, S2, S3, S4, and S5.

